# Predicting the onset of freezing of gait in de novo Parkinson’s disease

**DOI:** 10.1101/2021.03.11.21253192

**Authors:** Fengting Wang, Yixin Pan, Miao Zhang, Kejia Hu

## Abstract

Freezing of gait (FoG) is a debilitating symptom of Parkinson’s disease (PD) related to higher risks of falls and poor quality of life. In this study, we predicted the onset of FoG in PD patients using a battery of risk factors from patients enrolled in the Parkinson’s Progression Markers Initiative (PPMI) cohort. The endpoint was the presence of FoG, which was assessed every year during the five-year follow-up visit. Overall, 212 PD patients were included in analysis. Seventy patients (33.0%) developed FoG during the visit (pre-FoG group). Age, bradykinesia, TD/PIGD classification, fatigue, cognitive impairment, impaired autonomic functions and sleep disorder were found to be significantly different in patients from pre-FoG and non-FoG groups at baseline. The logistic regression model showed that motor factors such as TD/PIGD classification (OR = 2.67, 95% CI = 1.41-5.09), MDS-UPDRS part III score (OR = 1.05, 95% CI = 1.01-1.09) were associated with FoG occurrence. Several indicators representing non-motor symptoms such as SDMT total score (OR = 0.95, 95% CI = 0.91-0.98), HVLT immediate/Total recall (OR = 0.91, 95% CI = 0.86-0.97), MOCA (OR = 0.87, 95% CI = 0.76-0.99), Epworth Sleepiness Scale (OR = 1.13, 95% CI = 1.03-1.24), fatigue(OR = 1.98, 95% CI = 1.32-3.06), SCOPA-AUT gastrointestinal score (OR = 1.27, 95% CI = 1.09-1.49) and SCOPA-AUT urinary score (OR = 1.18, 95% CI = 1.06-1.32) were found to have the predictive value. PD patients that developed FoG showed a significant reduction of DAT uptake in the striatum. However, no difference at baseline was observed in genetic characteristics and CSF biomarkers between the two patient sets. Our model indicated that TD/PIGD classification, MDS-UPDRS total score, and Symbol Digit Modalities score were independent risk factors for the onset of FoG in PD patients. In conclusion, the combination of motor and non-motor features including the akinetic subtype and poor cognitive functions should be considered in identifying PD patients with high risks of FoG onset.

## Introduction

Freezing of gait (FoG), defined as a sudden inability to initiate or continue gait, is one of the most disabling symptoms in PD^1^. As a common symptom in advanced PD, it is associated with an increased risk of severe symptom, apathy and poor quality of life^2,3^. The episodic nature of FoG duration and a multifaceted pathophysiology make it hard to find a unified pathogenetic mechanism^4^. Yet studies indicate that the dopaminergic insult in the sensorimotor striatum, as well as dysfunctional cortical and cerebellar projections may contribute to its manifestation^1^. While specific therapy is still underway, early prediction of FoG could help identify FoG patients and take preventive measures beforehand^5^.

Previous studies have concentrated on complex determinants and pathophysiology of FoG^6^. Only a few studies have made efforts to predict FoG onset, especially in newly diagnosed PD patients^7-16^. Wearable sensors have been a heated topic used to detect and predict FoG, but these are applied to a limited number of patients and have a significant variability in outcomes measured^17^. Clinical variables, biomarkers and imaging examinations are feasible and convenient in evaluating FoG development. Several factors should be taken into consideration when identifying potential FoG patients. Motor factors such as higher scores of rigidity, postural instability, bradykinesia were predictive of FoG, as was shown in early studies^7^. The motor symptoms formed the akinetic-rigid PD subtype, which itself was identified to be a risk factor^8^. Non-motor factors included specific cognitive deficits, mood and sleep disorders, which were also found to be associated with FoG occurrence^8-12^. Imaging parameters included striatal dopaminergic denervation, which can be examined through the dopamine transporter (DAT) scans and single photon emission computed tomography (SPECT) imaging^6^. PD patients that had a lower DAT activity in caudate nucleus and putamen were at a higher risk to develop FoG^10^. Besides, white matter hyperintensities^13^, biomarkers in cerebral spinal fluid^9^ and gene mutations^14^ were suggested to be predictive of or involved in the mechanisms of FoG onset. While previous studies have helped identify several FoG risk factors, there were supporting and refuting evidence for the results. Most studies had the median follow up duration of less than five years, while several analyses have included PD patients with the presence of FoG symptom at baseline^7,15^. These were not consistent with our hope to identify PD patients at risk before symptom onset.

Instead of focusing on one certain aspect of clinical or imaging assessments, our study analyzed a comprehensive battery of indicators to predict FoG onset. Motor and non-motor factors, genetic characteristics, CSF biomarkers as well as the neuroimaging parameters were evaluated using longitudinal data of five-year visit from the PPMI cohort. Our goal was to determine the early symptoms and characteristics exhibited in PD patients before FoG occurrence and help elucidate the relations of these factors with FoG occurrence and development.

## Material and methods

### 1. Study design and participants

Data used in the preparation of this article were obtained from the PPMI database (www.ppmi-info.org/data). PPMI is a comprehensive observational, international, multi-center study designed to identify PD progression biomarkers. Its cohort comprises more than 400 recently diagnosed PD followed longitudinally for clinical, imaging and biospecimen biomarker assessment using standardized data acquisition protocols at 21 clinical sites. PD subjects were required to be over 30 years old, untreated with PD medications, within two years of diagnosis, Hoehn and Yahr < 3, and to have either at least two of resting tremor, bradykinesia, or rigidity (must have either resting tremor or bradykinesia) or a single asymmetric resting tremor or asymmetric bradykinesia at enrollment. Every study included in PPMI was approved by the institutional review board at each site and had obtained written informed consent provided by the participants.

Data from patients with PD with five-year follow-up were included in this analysis. The database was accessed on December 10, 2020. The last time for the update of the baseline data was April 20, 2020. Two patient groups formed in our study were based on the presence or absence of FoG within five years of visit. FoG was assessed using the reported value from Movement Disorders Society-Unified Parkinson’s Disease Rating Scale (MDS-UPDRS) Part II item ‘Freezing’ and Part III item ‘Freezing of gait’. The value scored of 1 or higher was converted to 1 representing the presence of FoG, while 0 represented no FoG onset. Overall, 212 subjects were included in the analysis with 142 non-FoG subjects and 70 subjects exhibiting FoG onset in five-year visit^18^.

### 2.2. Baseline assessments

A comprehensive collection of motor and non-motor features, as well as imaging assessments and genotyping, were recorded at baseline and each visit year. Motor symptoms included in the study were resting tremor, rigidity, bradykinesia, postural instability at diagnosis; Tremor Dominate (TD) /Postural Instability and Gait Difficulty (PIGD) classification; tremor score; Movement Disorders Society-Unified Parkinson Disease Rating Scale (MDS-UPDRS) part II and III. Modified Schwab & England ADL score was recorded to evaluate activities of daily living.

Non-motor symptoms were evaluated by the following items: MDS-UPDRS part I; Montreal Cognitive Assessment (MOCA) to assess global cognition; MCI test scores to evaluate test-based mild cognitive impairment (MCI); Hopkins Verbal Learning Test (HVLT) to assess memory; the 40-item University of Pennsylvania Smell Identification Test (UPSIT) to assess olfactory function; Benton Judgement of Line Orientation Test to assess visuospatial function; Epworth Sleepiness Scale and REM sleep behavior disorder questionnaire to assess sleep behavior; Geriatric Depression Scale, State-Trait Anxiety Inventory (STAI), and Questionnaire for Impulsive-Compulsive Disorders (QUIP) to assess neuron-behavior; Symbol Digit Modalities Test (SDMT) to assess processing speed-attention; Letter Number Sequencing and semantic (animal) fluency test to assess executive abilities-working memory; Scales for Outcomes in Parkinson’s Disease-Autonomic (SCOPA-AUT) to assess autonomic function.

Cerebrospinal fluid (CSF) biomarkers were recorded including CSF amyloid-β1-42 (Abeta), total (t)-tau (Tau), and phosphorylated tau (pTau) and alpha-synuclein (aSyn). Genetic patterns of the microtubule-associated protein tau (MAPT), the apolipoprotein ε4 (APOE4) allele, mutations in α-synuclein (SNCA) including SNCA_rs3910105 and SNCA_rs356181 were tested in the patient cohorts. Subjects with missing value in genetic patterns, imaging assessments and demographic information were screened out of the study.

### 2.3. SPECT-DAT and Structural MRI imaging

Indexes of reconstructed and attenuation-corrected 123I-FP-CIT SPECT imaging data was downloaded from the PPMI website. All participants underwent a DAT imaging procedure to measure the amount of dopamine in the brain using SPECT with the DAT tracer 123I-ioflupane at baseline. The striatal-binding ratios were calculated for left and right striatum separately using the occipital lobe as reference, thus obtaining ipsilateral, contralateral, and mean measurements. The asymmetry index was calculated as below:

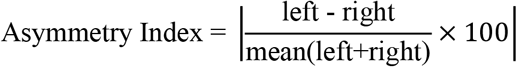

The count density ratio was calculated as below:

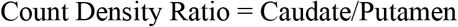

Seventy-eight patients included in our study had baseline gray matter volume recorded in MRI imaging given on the website. The value of gray matter volume was extracted from high resolution T1-weighted 3T MRI imaging. The gray matter volume was also included for imaging analysis.

### 2.4. Statistical analysis

R (v.4.0.1) was used for statistical analysis. All continuous data that followed the normal distribution examined by Shapiro-Wilks test were presented as the mean (the standard deviation). Continuous data that were not normally distributed were presented as median[quartile]. Student’s t-test, Kruskal Willis test, chi square and fisher exact test were used to compare baseline features between patients with and without FoG. *P* value < 0.05 was presented with ‘*’, < 0.01 was presented with ‘**’, < 0.001 was presented with ‘***’. Multivariate logistic regressions were conducted for factors that had *P* value < 0.05 in the initial univariate testing. Odds ratio (OR) and 95% confidence interval (CI) were reported for both the bivariate and the multivariate analyses. Indexes with *P* value < 0.05 in univariate analysis were put in the initial logistic regression model. The initial logistic regression model was built based on backward stepwise selection with the threshold of *P* value set as 0.05. The outcome variable was presence or absence of FoG within five years of follow up. A receiver operating characteristic (ROC) curve was created to assess sensitivity and specificity of the predictive model. The Hosmer and Lemeshow goodness-of-fit test was used to assess the model calibration.

## Results

### Baseline characteristics

Overall, 70 (33.0%) out of 212 PD patients had developed FoG symptoms within five years. Significant difference was observed in the age (*P* = 0.037*), age at symptom onset (*P* = 0.019*) and age at PD diagnosis (*P* = 0.031*) between patients that developed FoG withing five years (pre-FoG patients) and patients without FoG symptoms (non-FoG patients). No significant difference in genetic characteristics as well as CSF biomarkers was observed at the baseline (**Table 1**). Imaging analysis showed that both left and right, ipsilateral and contralateral caudate, putamen and the striatum had significantly decreased DAT uptake in pre-FoG patients compared with Non-FoG patients. The median time of the pre-FoG patients developing FoG was 2.61 years since diagnosis.

**Table 1.**
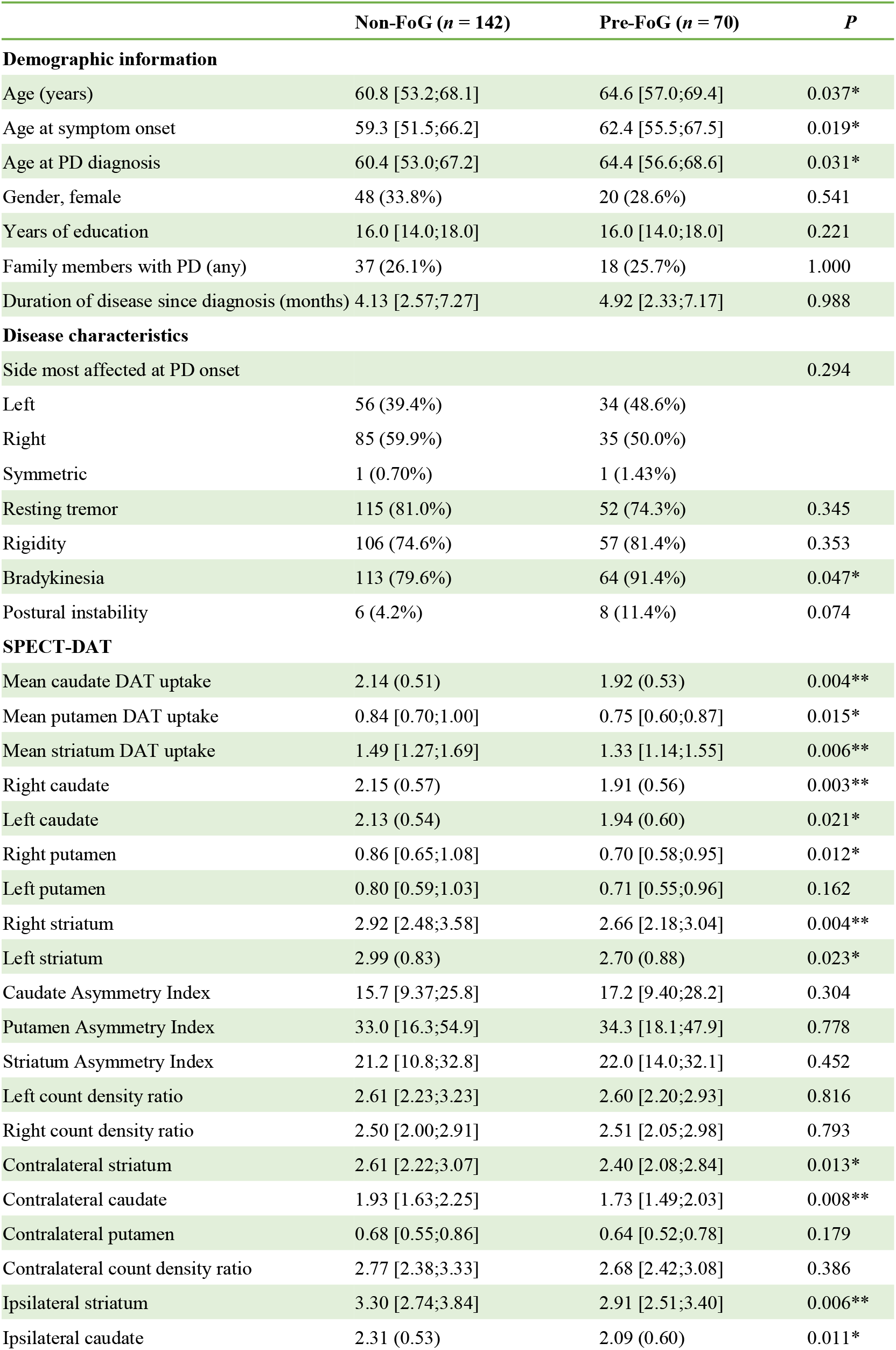
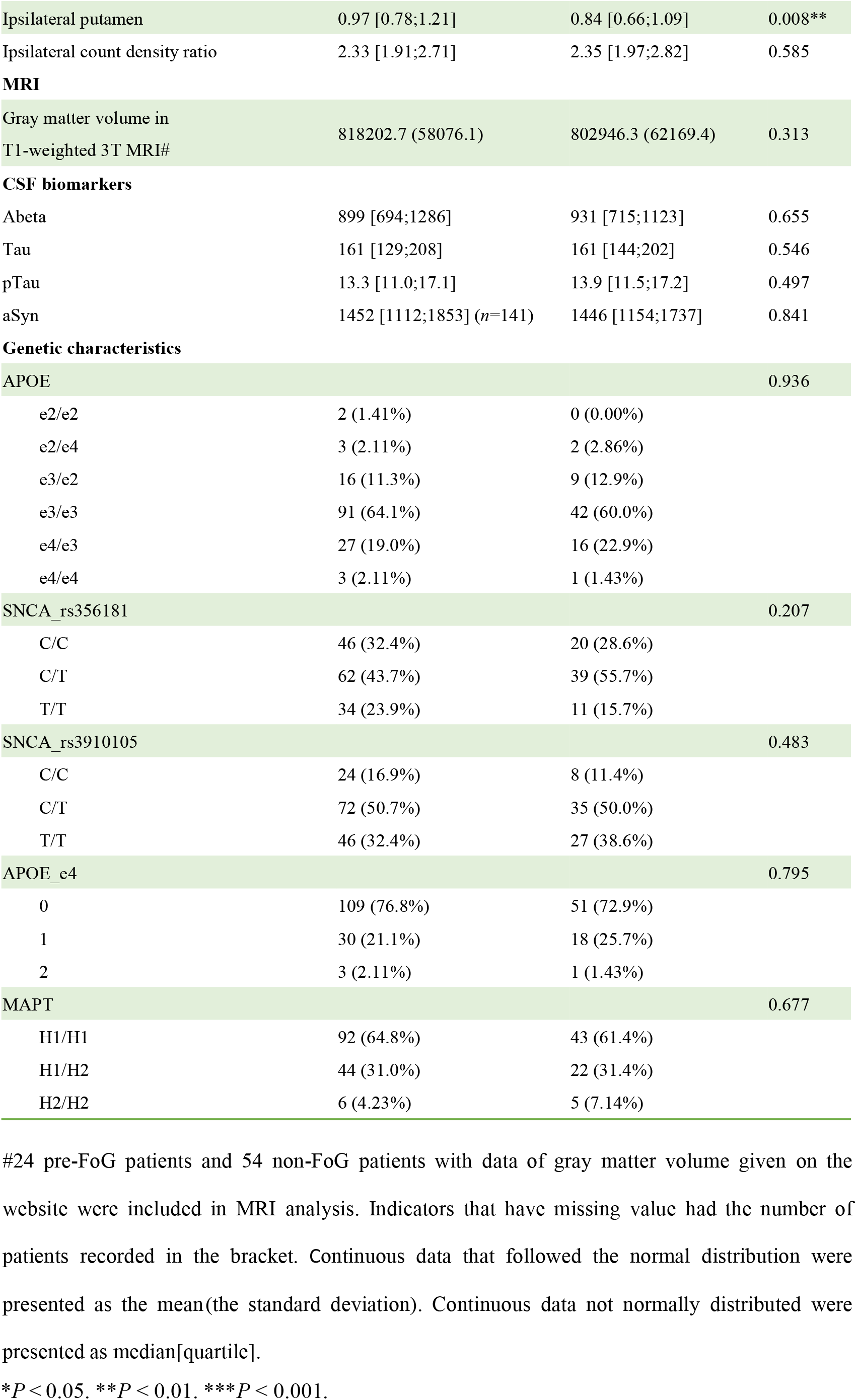
Demographic, disease, imaging and genetic characteristics of pre-FoG and non-FoG subjects with Parkinson’s disease at baseline.

In the motor and non-motor assessments of non-FoG and pre-FoG patients (**Table 2**), significant differences were observed in motor behavior such as TD/PIGD classification (*P* = 0.009**), MDS-UPDRS Part II/III Scores, bradykinesia (**Table 1**, *P* = 0.047*) as well as non-motor examinations such as SDMT Score (*P* < 0.001***), Epworth Sleepiness Scale Score (*P* = 0.005**), HVLT Immediate/Total Recall (*P* = 0.003**), SCOPA-AUT Gastrointestinal Score (*P* = 0.006**), SCOPA-AUT Urinary Score (*P* = 0.024**), SCOPA-AUT Cardiovascular Score (*P* = 0.048*), and MOCA Score (*P* = 0.039*). At the visit year five, apart from all the differences showed at baseline, more distinctions were exhibited in Categorical Hoehn & Yahr (*P* = 0.002**), Modified Schwab & England ADL Score (*P* < 0.001***), depressed mood (*P* = 0.011*), apathy (*P* = 0.044*), Benton Judgement of Line Orientation Score (*P* = 0.006**), Geriatric Depression Scale Score (*P* = 0.001**), HVLT Discrimination Recognition (*P* = 0.001**), Letter Number Sequencing Score (*P* = 0.006**), SCOPA-AUT Thermoregulatory Score (*P* = 0.023*), SCOPA-AUT Total Score (*P* < 0.001***), Semantic Fluency Total Score (*P* = 0.005**) and STAI Total Score (*P* = 0.015*).

**Table 2.**
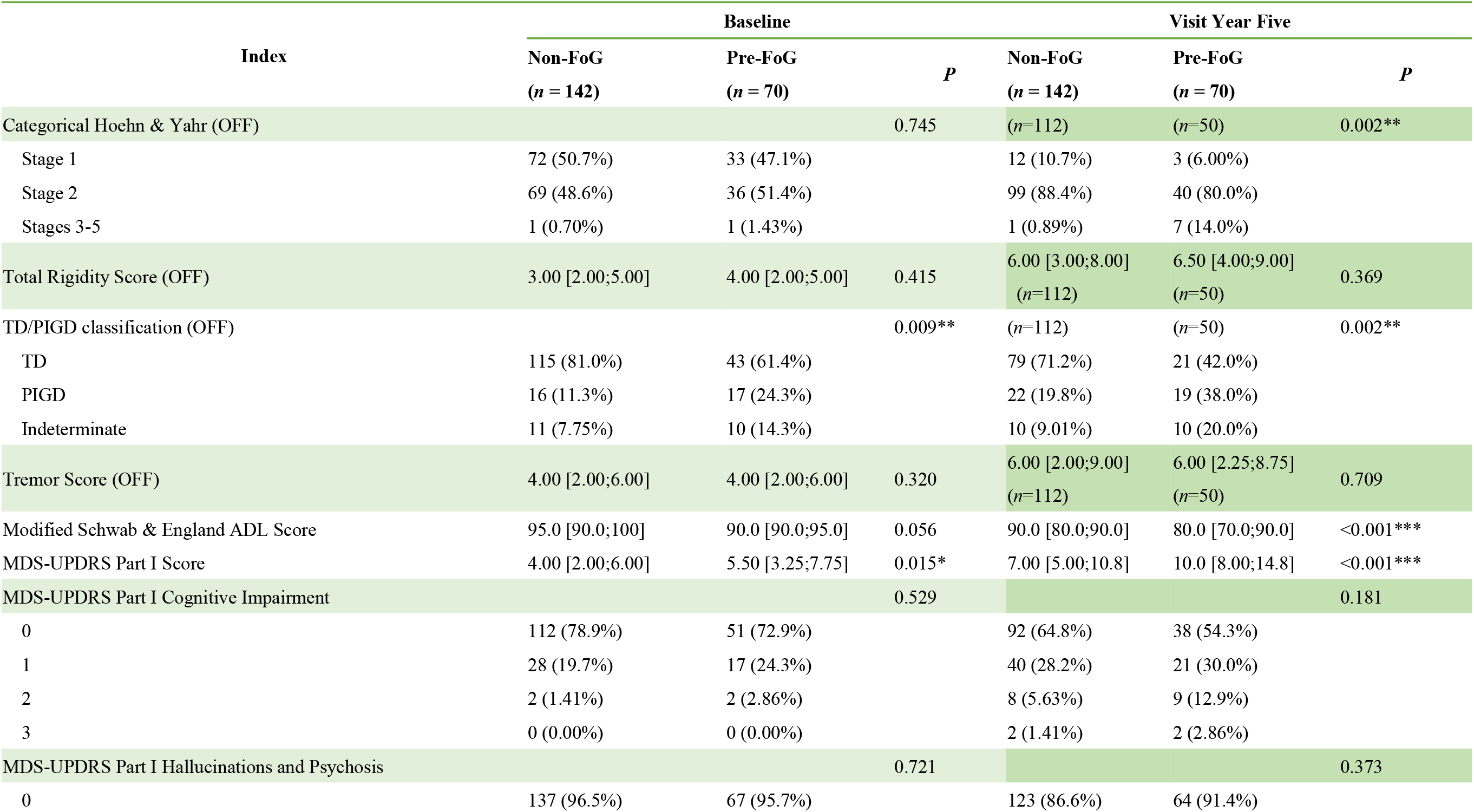
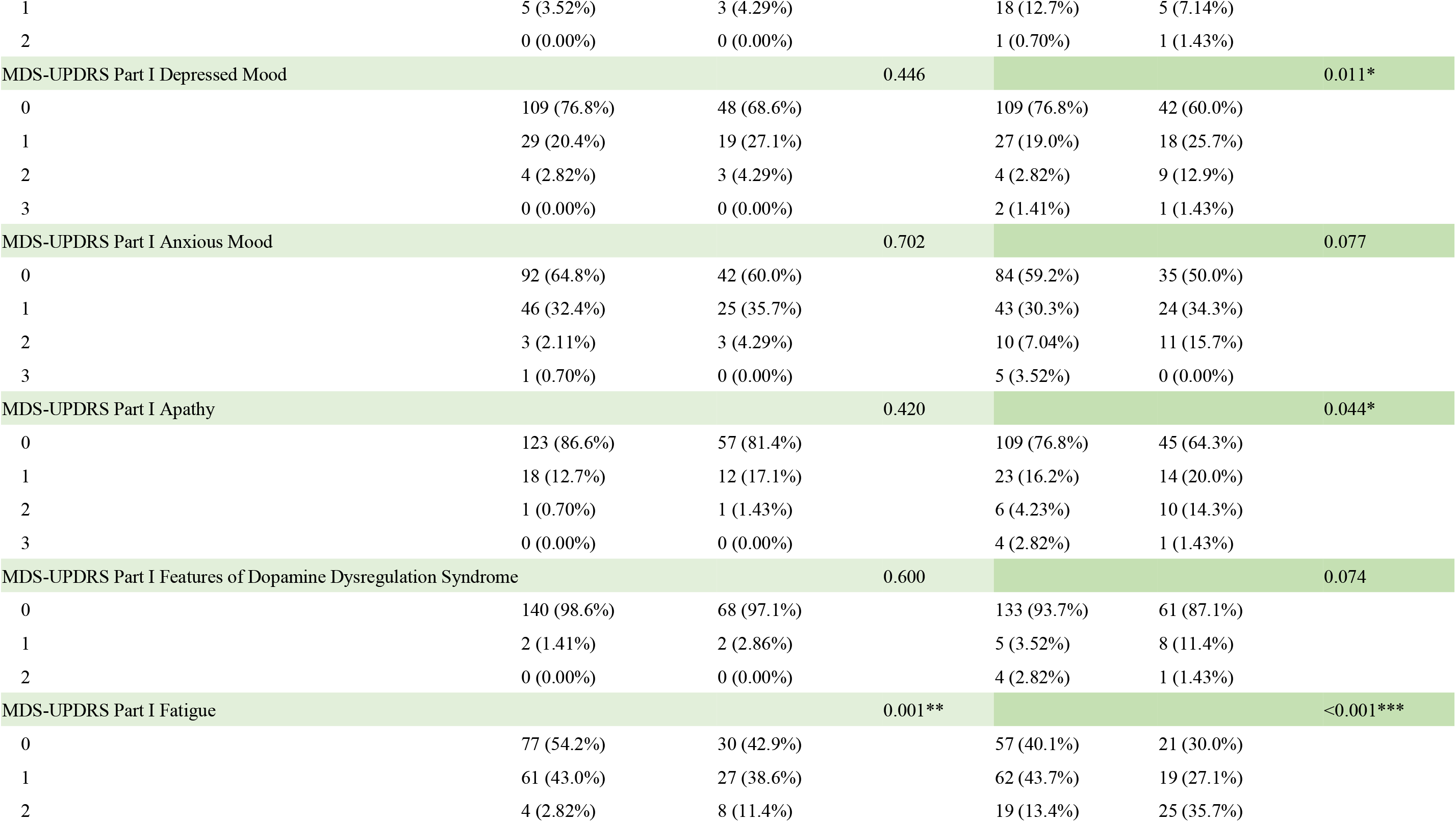
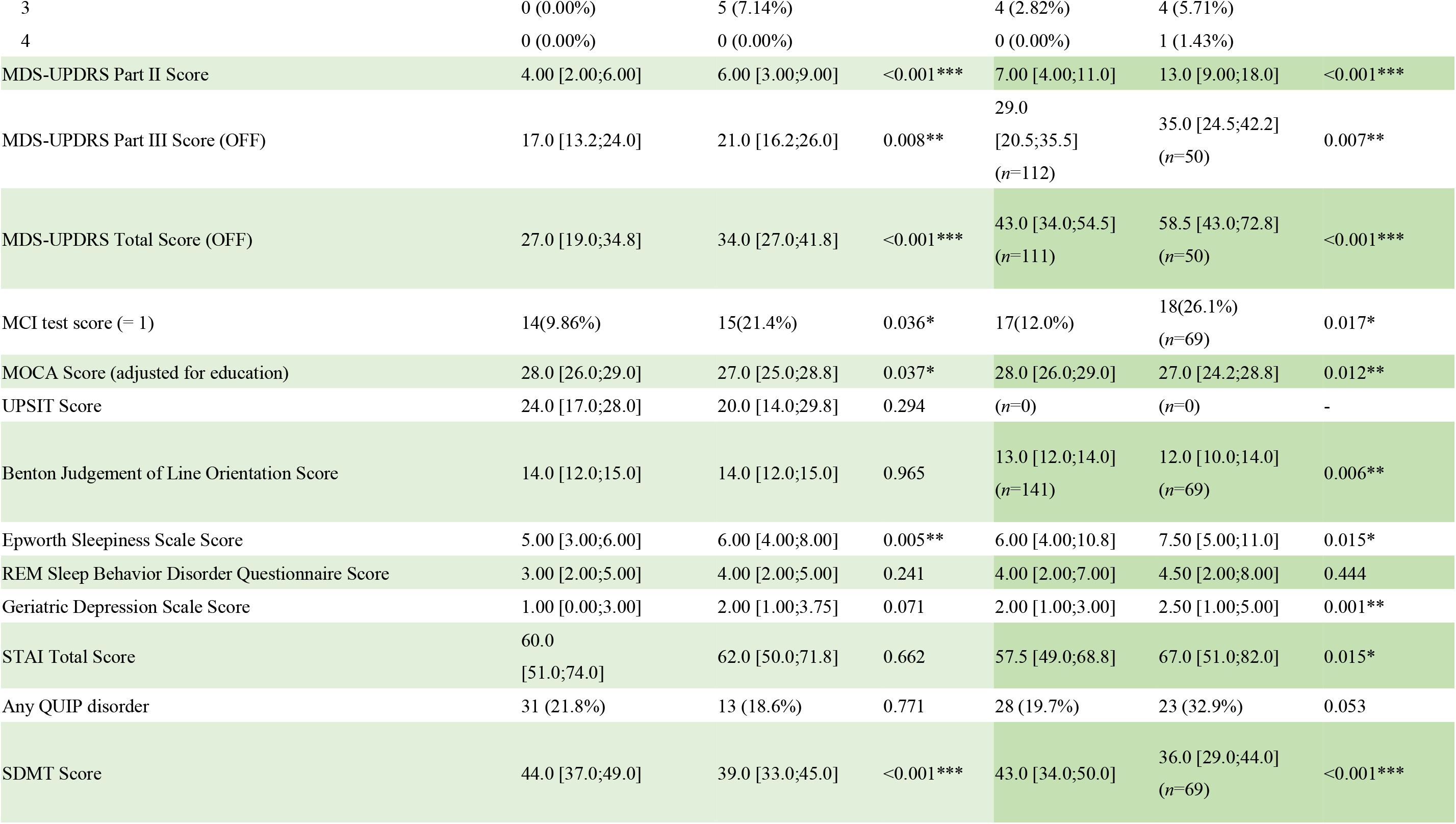
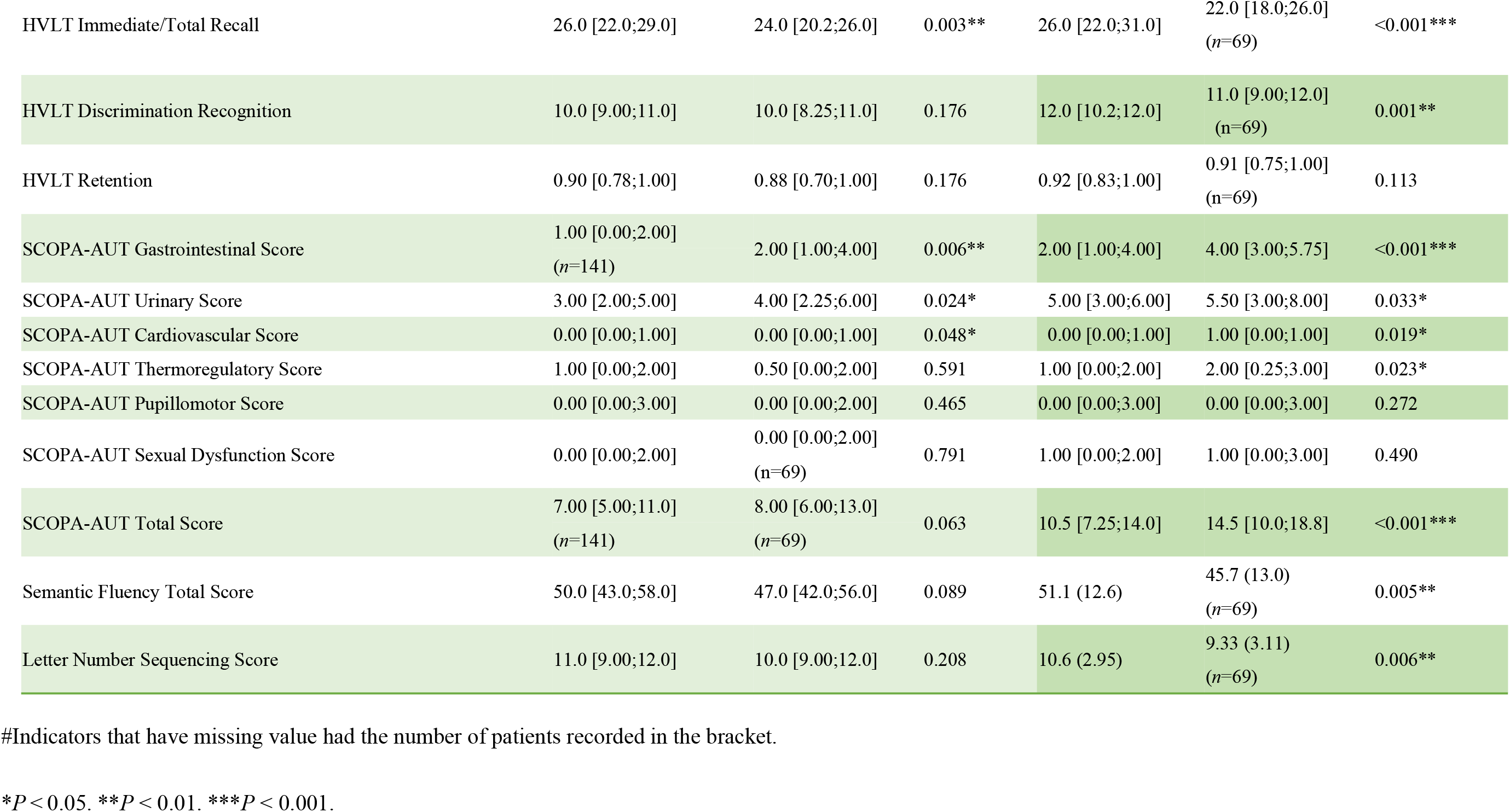
Motor and non-motor assessments of pre-FoG and non-FoG subjects with Parkinson’s disease at baseline and visit year five.

### Motor assessments

MDS-UPDRS part II score (OR = 1.13, 95% CI = 1.05-1.22), MDS-UPDRS part III score (OR = 1.05, 95% CI = 1.01-1.09), TD/ PIGD classification (OR = 2.67, 95% CI = 1.41-5.09) and bradykinesia (OR = 2.74, 95% CI = 1.15-7.62) were found to be correlated with FoG occurrence. Post hoc analyses revealed that non-FoG patients had a significantly higher TD/PIGD ratio compared with pre-FoG group (*P* = 0.009**). Freezers had significantly higher MDS-UPDRS part III score (*P* = 0.011*) compared with non-freezers, indicating worse motor examinations exhibited at baseline. No predictive power of other initial symptoms including resting tremor, rigidity and postural instability was observed between groups (**Table 3**).

**Table 3.** Univariate analysis of motor symptoms at baseline for FoG onset within 5 years of follow up.

| Index | OR | 95% CI |  | P |
| --- | --- | --- | --- | --- |
| Resting tremor | 0.68 | 0.35 | 1.35 | 0.263 |
| Rigidity | 1.49 | 0.75 | 3.12 | 0.273 |
| Bradykinesia | 2.74 | 1.15 | 7.62 | 0.034* |
| Postural instability | 2.92 | 0.98 | 9.23 | 0.056 |
| Total rigidity score | 1.03 | 0.92 | 1.15 | 0.607 |
| TD/PIGD classification (TD subtype) | 2.67 | 1.41 | 5.09 | 0.003** |
| Tremor | 0.94 | 0.85 | 1.04 | 0.244 |
| MDS-UPDRS part II score | 1.13 | 1.05 | 1.22 | 0.001** |
| MDS-UPDRS part III score | 1.05 | 1.01 | 1.09 | 0.011* |
| MDS-UPDRS total score | 1.05 | 1.02 | 1.07 | <0.001*** |
| Modified Schwab & England ADL score | 0.95 | 0.90 | 1.00 | 0.079 |

\* $P < 0.05$ . \*\* $P < 0.01$ . \*\*\* $P < 0.001$ .

### Non-Motor assessments

Several test scores were found to be related with FoG occurrence. Cognitive tests including SDMT (OR = 0.95, 95% CI = 0.91-0.98), HVLT immediate/Total recall (OR = 0.91, 95% CI = 0.86-0.97), MOCA (OR = 0.87, 95% CI = 0.76-0.99), MCI test score (OR = 2.49, 95% CI = 1.12-5.57) were found to have significant predicting power (**Table 4**).

**Table 4.** Univariate analysis of nonmotor symptoms at baseline for FoG onset within 5 years of follow up.

| Index | OR | 95% CI |  | P |
| --- | --- | --- | --- | --- |
| MCI test score (= 1) | 2.49 | 1.12 | 5.57 | 0.024* |
| MOCA score | 0.87 | 0.76 | 0.99 | 0.030* |
| MDS-UPDRS part I score | 1.12 | 1.03 | 1.23 | 0.007** |
| MDS-UPDRS Part I Anxious Mood | 1.18 | 0.72 | 1.93 | 0.506 |
| MDS-UPDRS Part I Apathy | 1.44 | 0.70 | 2.92 | 0.304 |
| MDS-UPDRS Part I Cognitive Impairment | 1.38 | 0.76 | 2.47 | 0.284 |
| MDS-UPDRS Part I Features of Dopamine Dysregulation Syndrome | 2.06 | 0.24 | 17.46 | 0.475 |
| MDS-UPDRS Part I Depressed Mood | 1.41 | 0.82 | 2.39 | 0.208 |
| MDS-UPDRS Part I Fatigue | 1.98 | 1.32 | 3.06 | 0.001** |
| MDS-UPDRS Part I Hallucinations and Psychosis | 1.23 | 0.25 | 5.15 | 0.784 |
| UPSIT score | 0.98 | 0.95 | 1.02 | 0.315 |
| Benton Judgement of Line Orientation score | 0.99 | 0.87 | 1.14 | 0.908 |
| REM sleep behavior disorder questionnaire score | 1.06 | 0.94 | 1.19 | 0.320 |
| Epworth Sleepiness Scale score | 1.13 | 1.03 | 1.24 | 0.012* |
| Geriatric Depression Scale score | 1.15 | 1.00 | 1.32 | 0.048* |
| STAI total score | 1.00 | 0.99 | 1.02 | 0.560 |
| QUIP score | 0.97 | 0.56 | 1.59 | 0.899 |
| SDMT score | 0.95 | 0.91 | 0.98 | 0.001** |
| HVLT immediate/Total recall | 0.91 | 0.86 | 0.97 | 0.004** |
| SCOPA-AUT total score | 1.07 | 1.02 | 1.12 | 0.009** |
| SCOPA-AUT cardiovascular score | 1.46 | 1.03 | 2.16 | 0.043* |
| SCOPA-AUT gastrointestinal score | 1.27 | 1.09 | 1.49 | 0.003** |
| SCOPA-AUT pupillomotor score | 0.77 | 0.46 | 1.22 | 0.285 |
| SCOPA-AUT sexual dysfunction score | 1.09 | 0.91 | 1.30 | 0.366 |
| SCOPA-AUT thermoregulatory score | 0.95 | 0.76 | 1.18 | 0.661 |
| SCOPA-AUT urinary score | 1.18 | 1.06 | 1.32 | 0.003** |
| Semantic Fluency total score | 0.97 | 0.95 | 1.00 | 0.055 |
| Letter Number Sequencing score | 0.92 | 0.82 | 1.02 | 0.127 |

\* $P < 0.05$ . \*\* $P < 0.01$ . \*\*\* $P < 0.001$ .

Non-cognitive tests that were associated included sleep disturbance: Epworth Sleepiness Scale (OR = 1.13, 95% CI = 1.03-1.24), mood disorder : MDS-UPDRS Part I Fatigue(OR = 1.98, 95% CI = 1.32-3.06) and autonomic dysfunction: SCOPA-AUT gastrointestinal score(OR = 1.27, 95% CI = 1.09-1.49), SCOPA-AUT urinary score (OR = 1.18, 95% CI = 1.06-1.32), SCOPA-AUT cardiovascular score (OR = 1.46, 95% CI = 1.03-2.16) as well as SCOPA-AUT total score (OR = 1.07, 95% CI = 1.02-1.12). In general, cognitive impairment, sleep disturbance, autonomic dysfunction and fatigue were predictive of FoG occurrence (**Table 4**).

While MDS-UPDRS scores were strong predictors of FoG onset, we further tested whether the changes of the scores were associated with FoG. After screening out of the patients with no MDS-UPDRS scores recorded in year one, 177 patients with 120 patients (67.8%) that had FoG symptoms within five years were included. Our results showed that the increase of MDS-UPDRS part II score (OR = 1.15, 95% CI = 1.05-1.26, *P* = 0.003**) and MDS-UPDRS total score (OR = 1.03, 95% CI = 1.00-1.07, *P* = 0.030*) from baseline to year-one were risk factors of FoG. However, changes in MDS-UPDRS part I score (*P* = 0.133) and MDS-UPDRS part III score (*P* = 0.337) were not associated with FoG onset in the study.

### DAT uptake and MRI analysis

Reduction of DAT uptake in the striatum, more specifically, in the caudate striatum (OR = 0.43, 95% CI = 0.23-0.76, *P* = 0.004**) was a strong predictive factor for FoG occurrence. However, the asymmetry index of striatum showed no predictive power in our analysis. In patient sets with gray matter volume extracted from T1-weighted 3T MRI imaging, the value did not present as an independent predictive factor (**Table 5**).

**Table 5.**
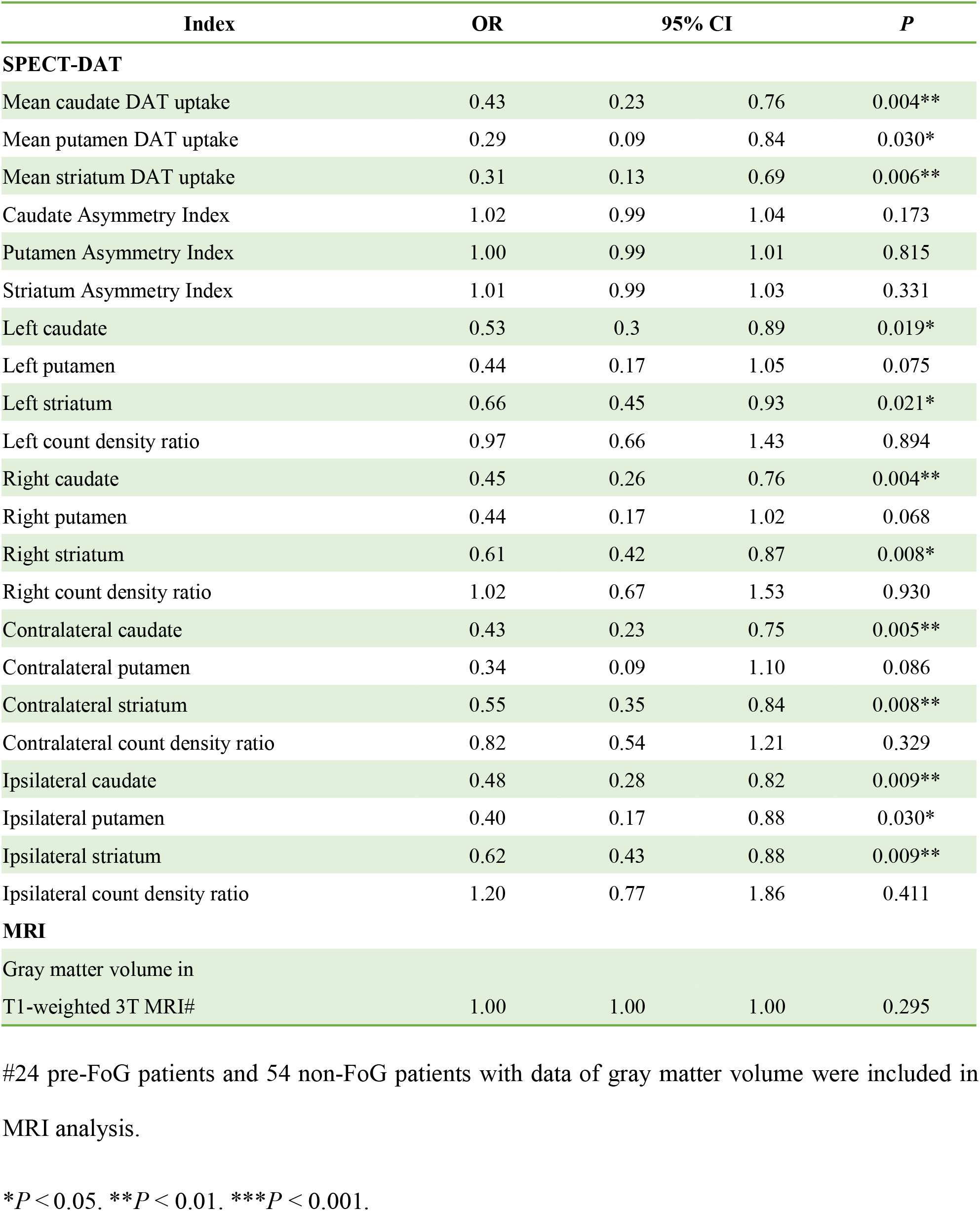
Univariate analysis of imaging data at baseline for FoG onset within 5 years of follow up.

None of the CSF biomarkers including amyloid-β 1-42, total Tau, phosphorylated Tau, total alpha-synuclein as well as genetic features including MAPT, the APOE4 allele, SNCA_rs3910105 and SNCA_rs356181 showed any predictive power in the univariate analysis. Factors that showed predictive value in univariate analysis were depicted in **Figure 1**.

**Figure 1.**
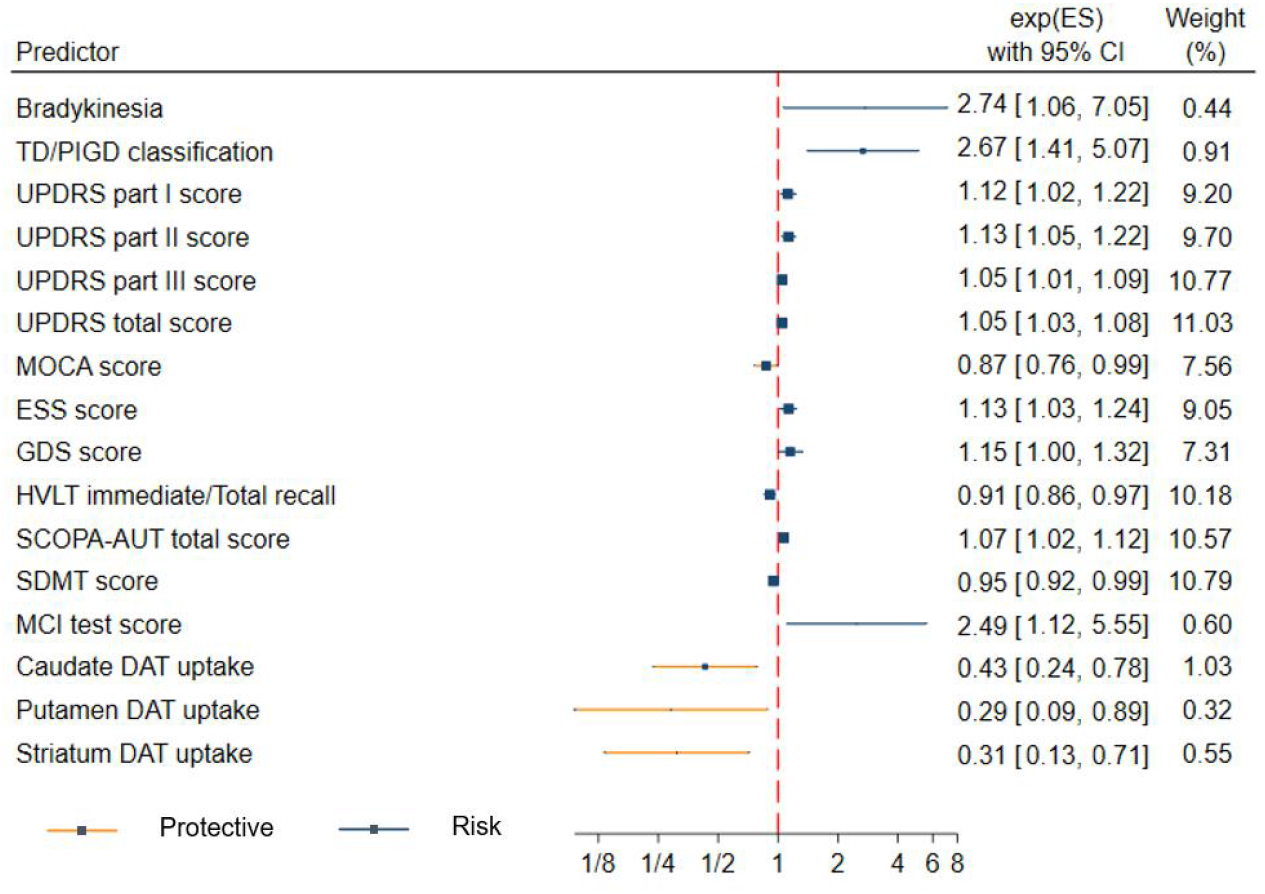
Predictors of the onset of FoG within 5 years follow up. Higher DAT uptake in striatum including caudate and putamen, better cognitive functions such as higher MOCA score, higher HVLT immediate/Total recall value and higher SDMT score (in red) are considered protective factors. Risk factors (in blue) include worse cognitive functions represented as higher MCI test score, worse autonomic functions, depression, sleep disturbance, PIGD subtype and bradykinesia.

### Predictive model of FoG

Three factors including TD/PIGD classification (OR = 2.87, *P* = 0.002**), MDS-UPDRS total score (OR = 1.04, *P* = 0.007**), and SDMT score (OR = 0.95, *P* = 0.008**) were selected from all the indicators with *P* value <0.05 at baseline in the multivariate analysis of the logistic regression. MDS-UPDRS total score was the sum of MDS-UPDRS Parts I, II, III, which included both motor examinations and non-motor tests. All the factors showed strong association with FoG occurrence in PD patients. PD patients with the PIGD subtype, higher MDS-UPDRS total score and lower SDMT score were at a higher risk of developing FoG (**Table 6)**. The area under curve (AUC) in the ROC analysis was 0.73 (**Figure 2**). The *P* value of the Hosmer and Lemeshow goodness-of-fit test was 0.598, indicating good calibrations.

**Table 6.** Multivariate analysis at baseline for the onset of FoG within 5 years of follow up.

| Index | OR | 95% CI |  | P |
| --- | --- | --- | --- | --- |
| TD/PIGD classification | 2.87 | 1.45 | 5.72 | 0.002** |
| MDS-UPDRS total score | 1.04 | 1.01 | 1.07 | 0.007** |
| SDMT score | 0.95 | 0.92 | 0.99 | 0.008** |

\* $P < 0.05$ . \*\* $P < 0.01$ . \*\*\* $P < 0.001$ .

**Figure 2.**
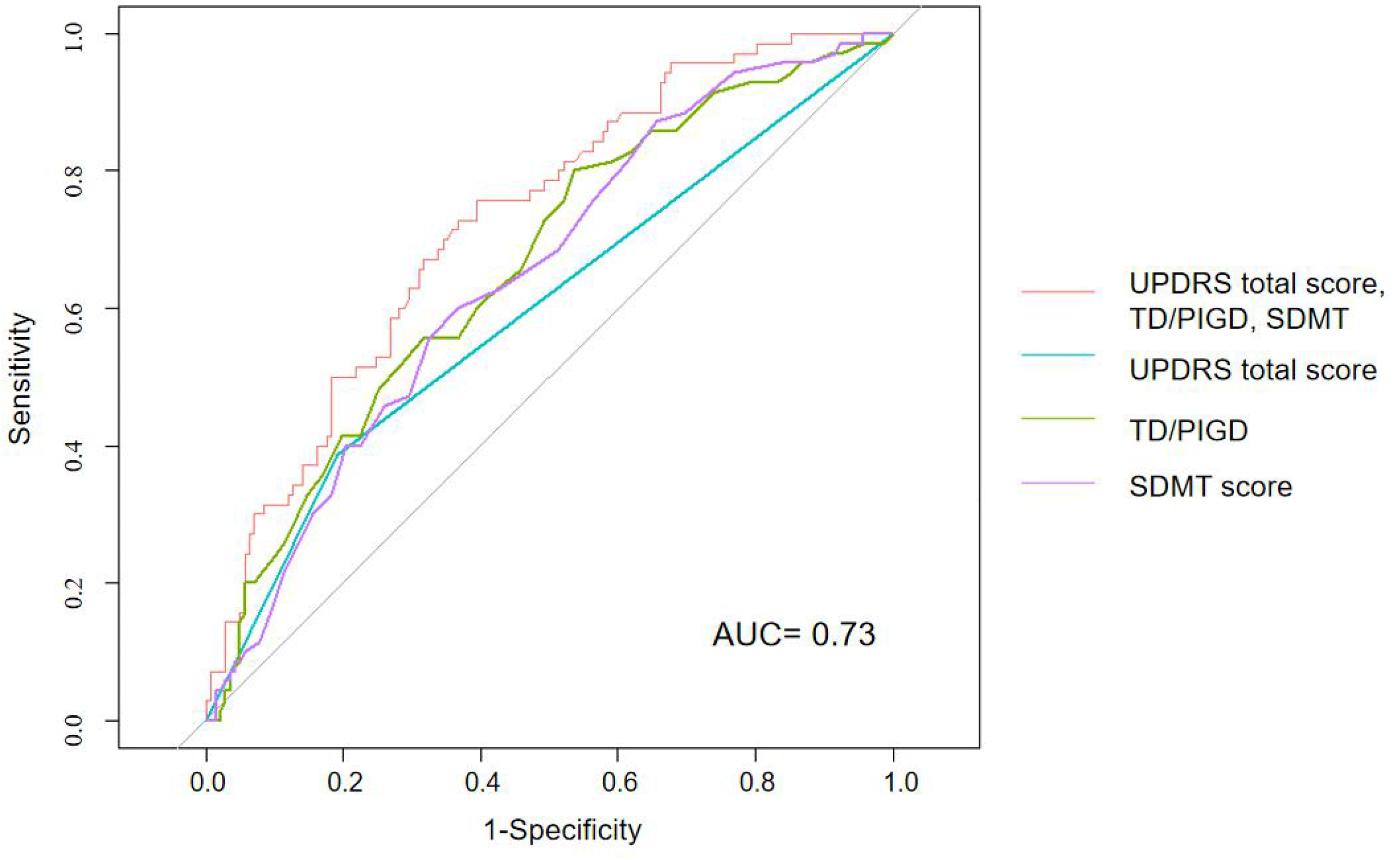
ROC curve analysis for the onset of FoG within 5 years follow up. The AUC for the multivariate predictive model (UPDRS total score, TD/PIGD and SDMT score) is 0.73.

The following equation represents the probability (*p*) of developing FOG in the five following years was presented as below:

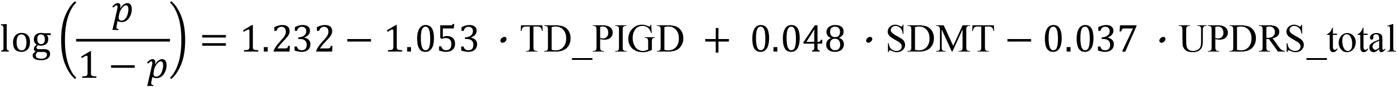

## Discussion

FoG had a significant negative impact on the health-related quality of life in PD patients^**19**^. Finding risk factors for FoG patients could help prevent potential outcomes and understand FoG mechanisms in the PD patients. Conflicting results regarding modifiable and non-modifiable risk factors for FoG occurrence were shown in the research before^6^. Our study tried to integrate various aspects of risk factors, using data from de novo PD patients in PPMI cohorts, to predict FoG onset in the early stage of PD. Excluding PD patients with FoG symptoms at baseline, we included 70 patients with FoG occurrence in the follow up duration of five years, and 142 patients with no FoG onset reported. Differences of motor and non-motor examinations were shown at baseline and visit year five. The predictive power of multiple baseline characteristics from clinical symptoms (e.g., bradykinesia, PIGD subtype, fatigue), to imaging parameters (e.g., reduced striatal DAT update) was shown in our study. Multiple logistic regression demonstrated the importance of TD/PIGD classification, MDS-UPDRS total score, and SDMT score in the progression to FoG in PD patients.

First of all, our study confirmed that motor, non-motor risk factors such as PIGD subtype (*P* = 0.003**), bradykinesia (*P* = 0.034*), cognitive impairment, sleep disturbance, autonomic disorders and fatigue (*P* = 0.001**) were associated with FoG occurrence. In line with other studies, PD patients with cognitive impairment represented as MCI (*P* = 0.024*), lower MOCA score (*P* = 0.030*), lower SDMT score (*P* = 0.001**), lower HVLT immediate/Total recall(*P* = 0.004**) were prone to develop FoG episodes later on^8,18^. Frontal-executive deficits, characterized by the slowed processing speed, attention and working memory as well as non-cued facilitated recall in prefrontal cortex (PFC), especially the dorsal-lateral part, were predominantly involved in the neuropsychology^20^. While the dopamine-mediated dysfunction of the associative circuit of the basal ganglia which disconnect the dorsal caudate and the PFC was contributory, the limbic and cortical spread of Lewy pathology, the β-amyloid and tau deposition and the presence of genes such as presence SNCA, APOE4, and MAPT H1 haplotype were all proposed to be contributory to the cognitive decline^21^. However, despite the significant cognitive impairment displayed in pre-FoG patients at baseline and visit year five, our studies showed no significant difference in the genetic features and CSF biomarkers between pre-FoG and non-FoG patients. This suggested that these factors contributed to the cognitive impairment in PD patients, especially pre-FoG patients only mildly.

Freezers had significantly sleep disturbances compared with the non-freezers^22^. Consistent with other studies, our study showed that sleep disorder represented as a higher Epworth Sleepiness Scale score (*P* = 0.012*) was predictive of FoG occurrence^16,22^. The pedunculopontine nucleus (PPN), a target for deep brain stimulation (DBS) for the treatment of parkinsonian gait disorders, may be involved in the association, as the same cells of large cholinergic neurons in PPN were involved in both arousal and motor control, confirmed in vitro, in vivo, and across species^22^. DBS in this region caused sleepiness or rapid transition into sleep, besides improving motor abilities in the FoG patients, further proving the association of sleep disorder with FoG around this region^23^.

Gastrointestinal and urinary autonomic symptoms often predate PD diagnosis by several years^24^. In our study, the impaired gastrointestinal and urinary functions were also associated with FoG occurrence as disease proceeded. This was also confirmed in other studies^9,11^. One hypothesis suggested that PD initiated in the autonomic nerve endings in the gastro-intestinal mucosa, which then spread through the vagal nerve from the gastrointestinal tract to the central nervous system^25^. The idea was supported by the common finding of phosphorylated α -synuclein inclusions initiated in hyperbranched, non-myelinated neuron terminals^26^. However, there were few studies that investigated the mechanism of the connections between the autonomic dysfunctions and FoG development. Only one study, assessing the heart rate changes in PD patients during FoG, provided links between impaired motor blockades and autonomic nervous system^27^. There were two studies that found no significant difference of autonomic functions between pre-FoG and non-FoG patients^28,29^. The sensitivity of the measurements may account for the inconsistency as both studies derived the autonomic symptom sub-score from items of the MDS-UPDRS parts I and II, while our study used a validated and more concrete measure, SCOPA-AUT to assess the autonomic dysfunctions^30^.

Fatigue was a common symptom in the course of PD development, In FoG patients, the symptom became even more prevalent^31^. As was confirmed in both our and other studies^12,32^, fatigue was also proved to be a risk factor for FoG development. Besides, the clinical feature was associated with many other non-motor symptoms, including daytime sleepiness. While dopaminergic neurons were mostly depleted in PD patients^6^, studies suggested that fatigue was most likely to result from disruption of nondopaminergic pathways^33^. Pavese N et al. found that the serotoninergic dysfunction in basal ganglia and limbic circuits was associated with fatigue, which also supported the observed associations between fatigue and other non-motor symptoms^33^. In relation with FoG, previous studies have found that the concentration of total serotonin had significant negative correlations with gait freezing^34^. Selective serotonin reuptake inhibitors (SSRIs) and serotonin and norepinephrine reuptake inhibitors (SNRIs) can improve FoG symptoms^35^, which further supported the serotoninergic association with FoG. However, the evidence supporting the anti-depressant therapy of FoG was limited. More studies are needed to validate the efficacy and the pathology of FoG with other nondopaminergic systems.

Second, our model demonstrated that the PIGD subtype (P = 0.002**), MDS-UPDRS total score (P = 0.007**) and SDMT score (P = 0.008**) were independent risk factors for FoG occurrence. PD patients experiencing freezing during the walks were more likely to be PIGD subtype^36^, suggesting preexisting gait impairment before FoG occurrence. The predisposing gait impairment provided evidence for the threshold model, which believed that the occurrence of FoG might be a joint effect of several gait disorders that accumulated to a threshold of impairment, finally leading to motor breakdown^6,37^. Since our studies excluded patients with freezing symptoms at baseline, there was no need to worry that the PIGD subtype evaluated by the sum of MDS-UPDRS items may self-include the ‘freezing’ itself, which was noticed in some other studies^38,39^. MDS-UPDRS total score was a composite score of both motor and non-motor examinations. While it stood out in our analysis, some studies showed that neither scale of MDS-UPDRS-I/II/III or the total score was well targeted to early PD patients^40^. Evaluating the change of scores using minimal but clinically relevant differences^41^, instead of comparing the baseline difference directly, may be a better examination in defining the disease severity and identifying FoG patients. In our study, we briefly evaluated the increase in MDS-UPDRS-I and MDS-UPDRS total scores after one year from baseline evaluation and confirmed the association, suggesting the feasibility of the methods. SDMT is a commonly used instrument to measure information processing speed, which is required in many cognitive operations^42^. It is sensitive to multiple neurological disorders, including multiple sclerosis, stroke and PD^42^. While SDMT task performance was a mixed result of visual and motor confounding factors in addition to cognition, a recent study using gaze analysis technique showed that the impaired performance of PD-MCI patients in the written SDMT task were not, even in part, due to visuomotor impairments, further emphasizing the role of cognitive functions played in PD patients in the test. However, since current studies are more concentrated on the relations of SDMT with multiple sclerosis, the physiologic and clinical relations of SDMT performance with FoG remains to be investigated^43,44^.

In line with other studies, our results showed that the more severe loss of striatal dopaminergic binding, shown in early stage of PD patients, could help predict FoG occurrence. While presynaptic striatal dopaminergic denervation was identified as a risk factor before^10^, our study further elucidated that the denervation appears in both left and right, contralateral and ipsilateral striatum, especially in the caudate nucleus. Despite the heterogeneity of neuronal mechanisms in FoG development, anti-coupling between cognitive cortical regions and the caudate nucleus was an independent factor of symptom severity, representing common neural underpinnings of freezing^45^. While evidence suggested the role of hemispheric laterality played in the development of freezing, no difference in the side of striatum degeneration, indicated by the count density ratio in our analysis was observed in our study^46^.

There were also some negative results shown in our studies. While we had taken genetic characteristics in consideration, our study failed to disclose any predictive power in the items selected. The APOE ε4 allele was previously reported to be a genetic risk factor for FoG in PD^47^. This association seemed to be mediated by Aβ-dependent pathways, which itself can independently contribute to the onset of FoG^47^. However, there was controversy regarding the predictive value of the APOE ε4 allele, as another study conducted by Factor et al. showed that the APOE ε4 allele was not associated with FoG in PD patients, and even decreased the risk of postural instability with falling^48^. While SNCA and MAPT genes have been consistently associated with the risk of developing PD^49^, these genes tend to relate more with the non-motor functions^50^. Potential confounders such as protective genes (CYP2D6*4, LRRK2 and different freezing phenotype may interfere with our analysis^36,48,51^. In our study, gray matter volume was not considered a risk factor for FoG occurrence within five years. There were also studies showing no significant difference between FoG+ and FoG-patients in terms of structural gray matter changes^52,53^. More investigations in the specific pattern of gray matter (GM) tissue were recommended to help identify the difference^54^.

Our studies also have several limitations. First, it is a retrospective study with 212 patients included in a follow-up range of five years. Limited number and of patients included as well as a relatively short period of time may not fully reveal the impact of risk factors in PD patients. Second, while numerous studies have evaluated FoG presence based on self-reported questionnaire, the method can easily mix FoG with other symptoms such as off-state akinesia, given the similar locomotive features. Use of quantitative objective FoG assessments or FoG diagnosis confirmed from wearable sensors may improve the accuracy. Third, selection bias should be noticed as we fully depended on data from PPMI and screened out the patients with no complete data. Other factors influencing the enrollment of subjects or the loss of information in these patients may be neglected in our study. More scrutinized selection of the patients and the matching of patient groups based on different symptoms can further elucidate the risk factors in FoG patients.

In summary, our findings determined the risk factors of FoG occurrence among a battery of clinical, imaging as well as genetic characteristics. Our results emphasized the importance of several factors in predicting FoG onset, including PIGD subtype, high MDS-UPDRS total score as risk factors and high SDMT score as a protective factor. Combining these results with further studies in randomized clinical trials and physiological research will help find early therapeutic targets to prevent or postpone FoG onset.

## Data Availability

Data used in the preparation of this article were obtained from the PPMI database (www.ppmi-info.org/data).

## Acknowledgements

This work was supported by Shanghai Pujiang Program (KH, 19PJ1407500), Medical and Engineering Cross Research Fund from Shanghai Jiao Tong University (KH, YG2019QNA31), Shanghai Municipal Health Commission Clinical Study Special Fund (KH, 20194Y0067), Ruijin Hospital Guangci Excellence Youth Training Program (K.H, GCQN-2019-B10); Ruijin Youth NSFC Cultivation Fund (K.H, 2019QNPY01031). PPMI is sponsored and partially funded by The Michael J. Fox Foundation for Parkinson’s Research (MJFF).

## Author contributions

Kejia Hu designed and conceptualized the study. Fengting Wang collected the data, Fengting Wang and Kejia Hu analyzed and interpreted the data. Fengting Wang and Kejia Hu wrote the manuscript. Fengting Wang, Yixin Pan, Weiqiu Jin, Miao Zhang and Kejia Hu provided critical revisions that were important for the intellectual content. All authors read and approved the final version of the manuscript.

